# Rigorous hepatitis B surface antigen analyses and identification of hepatitis B chronicity amongst South Africans attending public health facilities over a five-year period: 2015 to 2019

**DOI:** 10.1101/2022.08.09.22278594

**Authors:** Shelina Moonsamy, Pavitra Pillay, Nishi Prabdial-Sing

**Affiliations:** Centre for Vaccines and Immunology, National Institute for Communicable Diseases, division of the National Health Laboratory Service, Johannesburg, South Africa; Department of Biomedical and Clinical Technology, Faculty of Health Sciences, Durban University of Technology, Durban, South Africa; Department of Medical Virology, School of Pathology, Faculty of Health Sciences, University of the Witwatersrand, Johannesburg, South Africa

## Abstract

Hepatitis B, a potentially life-threatening viral infection of the liver, remains a global public health concern despite the availability of effective vaccines for over three decades. Given that most HBsAg studies targeted distinct cohorts, we aimed to provide HBsAg data nationally in the public health sector of South Africa.

We conducted a cross-sectional study on HBsAg tests obtained from the National Health Laboratory Service Central Data Warehouse for tests performed nationally during the period 2015 to 2019. Annual data were cleaned and appended prior to data interrogation to determine and analyse the total number of cases who tested positive for HBsAg and the number of chronic HBV infections.

We identified 176,530 cases who tested positive for HBsAg at least once during the 5-year period, with a test positivity rate of 9%. Chronic infections were identified in 6.4% of HBsAg positive cases. Clearance of HBsAg was observed in 5,571 cases, inclusive of clearance in 135 chronic cases. Significantly more males tested positive for HBsAg and were chronically infected (p < 0.0001). Amongst individuals who were vaccine-eligible as infants (0 to 19 years old), 4,980 tested HBsAg positive, of which 22.7% (1,131) were under 5 years old, with a HBsAg population positivity rate of 0.02% and test positivity rate of 4.83%.

HBsAg positivity amongst vaccine-eligible individuals is likely due to suboptimal vaccine coverage rates reported for South Africa. Without a birth dose of the HBV vaccine and lack of routine HBsAg screening at antenatal care, it is likely that the majority of HBsAg positive cases under 5 years old were vertically infected. Optimal HBV vaccine coverage, inclusive of a birth dose, is key to eliminating horizontal and vertical transmission of HBV. Early identification of HBV chronicity is fundamental in reducing the risk of liver cirrhosis and hepatocellular carcinoma.

## 1. Background

Hepatitis B, a potentially life-threatening viral infection of the liver, remains a global public health concern despite the availability of effective vaccines for over three decades (1-3). It occurs following infection with the hepatitis B virus (HBV). HBV is transmitted through contact with blood or other body fluids of an infected person. Common transmission routes are sexual contact, intravenous drug use and vertical transmission from mother to child during birth (4). Infection with HBV can cause acute and chronic disease. The risk of progression to chronicity decreases with age, ranging from around 70-90% amongst infants less than a year old to 1-10% amongst individuals over 20 years old (5-7). Resolved HBV infection usually results in hepatitis B immunity that prevents disease following re-exposure, however HBV reactivation or reinfection, as identified by the reappearance of detectable HBsAg levels, has been reported in individuals infected with HIV-1 (8). Chronically infected individuals have a 15-40% lifetime risk of cirrhosis, liver failure or hepatocellular carcinoma and although chronic HBV infection has no known cure, disease progression can be managed with various treatment options as highlighted in the National Guidelines for the Management of Viral Hepatitis (9-11). In 2019, The World Health Organisation (WHO) estimated that 296 million people were living with chronic HBV infection, a 15% increase from the 2017 estimate of 257 million (12, 13). In June 2016, the World Health Assembly adopted a resolution to eliminate viral hepatitis as a major public health problem by 2030 (1). Elimination is targeted at reducing new chronic infections by 90% and HBV related deaths by 65% (1). In addition, one of the WHO impact targets for measuring elimination is ≤0.1% HBsAg prevalence in children under 5 years old (14).

HBV infection is diagnosed following laboratory detection of HBsAg, which indicates active infection (15). If HBsAg is not cleared within six months of first detection, HBV infection is classified as chronic (1, 16, 17). HBsAg clearance amongst chronically infected HBV individuals may occur in around 1-2% of individuals, either naturally or following treatment interventions (18, 19).

Hepatitis B was highly endemic in South Africa before April 1995 when the HBV vaccine was introduced as a monovalent dose into the South African expanded programme on immunisation (EPI) schedule (20). Since December 2015, the HBV vaccine is one of the vaccines amongst the hexavalent combination vaccine of diphtheria, tetanus, acellular pertussis, inactivated poliovirus, Haemophilus influenza type B and hepatitis B (DTaP-IPV-HIB-HepB). The current HBV vaccination schedule consists of doses at 6, 10 and 14 weeks of age, and a booster dose at 18 months of age (21, 22). Although recommended to minimise the risk of mother to child transmission of HBV, a birth dose of the HBV vaccine is yet to be included in the EPI schedule of South Africa (23, 24). Vaccination coverage for South Africa amongst children under one year old who have completed their primary course of immunisation has been reported from 2000 to 2019 by the Health Systems Trust of South Africa using data obtained from the District Health Information System (25-29). Over the 20-year period, the average coverage rates by province ranged from 67.2% in North West to 85.6% in Northern Cape (S1 Table).

In South Africa, chronic HBV prevalence has been estimated at 2.5 million cases, reported in 2014 (6). Based on the population size of South Africa in 2014 at just over 54 million, this would translate to a HBsAg prevalence of 4.6%, indicative of intermediate HBV endemicity (2 to 7% HBsAg prevalence) (20, 30-32). A major transmission route in South Africa was reported to be unexplainable horizontal transmission in children under 5 years of age, whilst sexual transmission was reported to be the major transmission route amongst adolescents and young adults (20).

Over the five years, 2015 to 2019, the average population of South Africa, as reported by Statistics South Africa was just under 57 million (Table 1) (33). Overall, there were higher numbers of females than males, although in Gauteng and North West, there were slightly more males than females. Provincially, the smallest province of South Africa (Gauteng), housed the majority of South Africans, whilst the largest province (Northern Cape, just over 20-fold larger) housed the least number of South Africans at more than 10-fold lower than Gauteng (Table 1).

**Table 1:** Land size versus population size by province of South Africa.

| Province | Land size |  | Population size |  |
| --- | --- | --- | --- | --- |
|  | Area (km<sup>2</sup>) (34) | Percentage | Number* (33) | Percentage |
| Northern Cape | 372,889 | 30.5 | 1,241,454 | 2.2 |
| Eastern Cape | 168,966 | 13.8 | 6,663,741 | 11.7 |
| Free State | 129,825 | 10.6 | 2,865,681 | 5.0 |
| Western Cape | 129,462 | 10.6 | 6,608,058 | 11.6 |
| Limpopo | 125,755 | 10.3 | 5,719,236 | 10.1 |
| North West | 104,882 | 8.7 | 3,893,193 | 6.8 |
| KwaZulu-Natal | 94,361 | 7.7 | 11,011,040 | 19.4 |
| Mpumalanga | 76,495 | 6.3 | 4,424,866 | 7.8 |
| Gauteng | 18,178 | 1.4 | 14,426,736 | 25.4 |
| **Total** | **1,220,813** | **100** | **56,854,006** | **100** |

\*The population size indicated here is the average population size from 2015 to 2019

Most HBsAg studies that have been conducted in South Africa targeted distinct cohorts (20, 22, 23, 35-40). Hence, the aim of the study was to describe and analyse the number of cases that tested positive, at least once, for HBsAg in a consecutive five-year period and to identify and analyse HBV chronic cases nationally in the public health sector of South Africa.

## 2. Methods

### 2.1 Study Population and Data Source

We conducted a cross-sectional study on HBsAg tests obtained from the National Health Laboratory Service (NHLS) Central Data Warehouse (CDW) for tests performed nationally during the period 2015 to 2019. The NHLS CDW is the national repository for laboratory data from the public health sector of South Africa, serving over 80% of the population (41). Variables of the dataset used for the purposes of this study were gender, date of birth (DOB), province (assigned to the testing facility), date of sample collection, name of testing facility, testing facility reference number (hospital identification number) and a CDW allocated unique identification number (UID). The UID is duplicated if a link to an existing record is identified based on the surname, first name and date of birth or national identification number, otherwise a new UID is generated. Criteria for inclusion in our study were negative and positive HBsAg results.

### 2.2 Data Cleaning and Appending

Data were initially cleaned per year from 2015 to 2019 in Microsoft Excel (Version 2016, Washington, USA) by identification, confirmation and deletion of quality assurance (non-patient related) records. Annual data were subsequently cleaned following importation into Stata/IC (Version 14.1, Texas, USA) to remove records of research study participants and format date variables according to the “DD/MM/YYYY” format prior to saving as Stata (.dta) files. Annual .dta files were appended into a single dataset containing all HBsAg test records from 2015 to 2019, sorted by the sample collection date and saved for further data manipulation.

### 2.3 Data Analyses

Data analyses were performed in in Stata/IC (Version 14.1, Texas, USA) and Microsoft Excel (Version 16, USA).

To determine the total number of cases who were tested for HBsAg over the five years, we de-duplicated the appended 2015 to 2019 dataset to obtain a line list of cases and used this as our denominator for HBsAg prevalence (%). To obtain a dataset of all HBsAg positive and negative records, we used the original appended dataset to identify and delete HBsAg records of equivocal results (neither positive nor negative) and sorted the dataset by the sample collection date (herein referred to as the master HBsAg dataset). We proceeded to identify cases that had only a single HBsAg test and cases with multiple HBsAg tests during 2015 to 2019. For those cases with more than one test, we merged the dataset so that a case would appear as a single line item with the variables of HBsAg test results and sample collection dates merged to include all results and sample collection dates of a specific case respectively. We then analysed the merged dataset over the five-year period to identify cases who tested negative throughout (Group_N), cases who tested positive throughout (Group_P), and cases who had mixed results of both negative and positive (Group_M). Group M was further interrogated to identify the number of cases that (1) tested negative after testing positive, (2) tested positive after an initial negative result and (3) tested negative in between two positive results.

#### 2.3.1 Development of a registry of cases who tested positive for HBsAg, 2015 to 2019

To identify cases that tested positive at least once over the 5 years, we selected all HBsAg positive records (comprising cases from Group_M and_Group P) and maintained a dataset of these records (herein referred to as the positive HBsAg positive dataset). As a registry requires a line list of cases, we de-duplicated the positive HBsAg dataset to delete all duplicate records of a case over the 5-year period to maintain a line list (registry) of cases.

#### 2.3.2 Identification of chronic HBV cases

We used the positive HBsAg dataset to identify and delete cases who had only one positive test over the five years, maintaining records of all cases who had more than one HBsAg positive test. We then merged the records to have each case appear as a single line item inclusive of all sample collection dates in ascending chronological order. We calculated the time interval in days between the first sample collection date and subsequent sample collection dates and maintained only records of cases who met the criteria of a time interval of 180 days (6 months) or more and classified them as chronic HBV cases (Group_C).

We subsequently determined how many chronic HBV cases were from Group_P and the different categories of Group_M by merging the Croup_C .dta file with Group_P and the different categories of Group_M .dta files using the UID as the key variable.

#### 2.3.3 Age group analysis

We calculated the age of each case based on the sample collection date and date of birth and subsequently assigned the cases to defined five-year age groups. With the master HBsAg dataset sorted by the sample collection date, age groups presented are based on the earliest sample collection date if a case had more than one record. We analysed age group 0 to 4 years old further to determine HBsAg test positivity rate (HBsAg positive cases / total cases 0 to 4 years old tested for HBsAg) and HBsAg population positivity rate (HBsAg positive cases / average population size 0 to 4 years old 2015 to 2019).

#### 2.3.4 Statistical Analyses

We analysed the data to determine the distribution of cases by age group and province stratified by gender, and to determine the number of HBsAg positive cases and HBV chronic cases per 100,000 province population. We used the average population over the five years (2015 to 2019) as reported by Statistics South Africa (33). To analyse statistical differences between gender by age group, we used Pearson’s chi-squared test in Stata/IC (Version 14.1, Texas, USA).

#### 2.3.5 Ethics

Ethics approval was obtained from the Faculty of Health Sciences, Institutional Research Ethics Committee (IREC 069/20) of the Durban University of Technology, Durban, South Africa. Approval to obtain the NHLS CDW data was obtained via the NHLS Academic Affairs and Research Management System (PR20254).

## 3. Results

Following data cleaning and appending of annual HBV data files, the appended 2015 to 2019 HBsAg dataset consisted of a total of 2,370,723 records (42). Deduplication of the dataset resulted in a line list of 1,957,224 cases (denominator for HBsAg prevalence) who were tested for HBsAg over the five years (Fig 1a). Of the 2,370,723 records, 6,211 records with equivocal results and were deleted, leaving 2,364,512 records of positive and negative HBsAg results (master HBsAg dataset) (Fig 1a). Of the master HBsAg dataset, 1,655,484 cases had only one HBsAg test conducted from 2015 to 2019 (Fig 1b). The remaining (709,028) were from cases who were tested for HBsAg at least twice during the five-year period. On merging the duplicates of the dataset of 709,028 records, we identified 297,267 cases with duplicate records; 252,429 cases in Group_N, 8,645 cases in Group_M and 36,193 cases in Group_P (Fig 1b). Of the 8,645 cases in Group_M, 5,571 cases tested negative after a positive test, 3,001 cases tested positive after a negative test, and 73 cases tested negative in between an initial and last positive test (Fig 1b). Of the 5,571 cases who tested negative after a positive test, 4,907 cases had tested positive initially and negative subsequently and 664 cases tested positive in between an initial and last negative test (Fig 1b).

**Figure 1:**
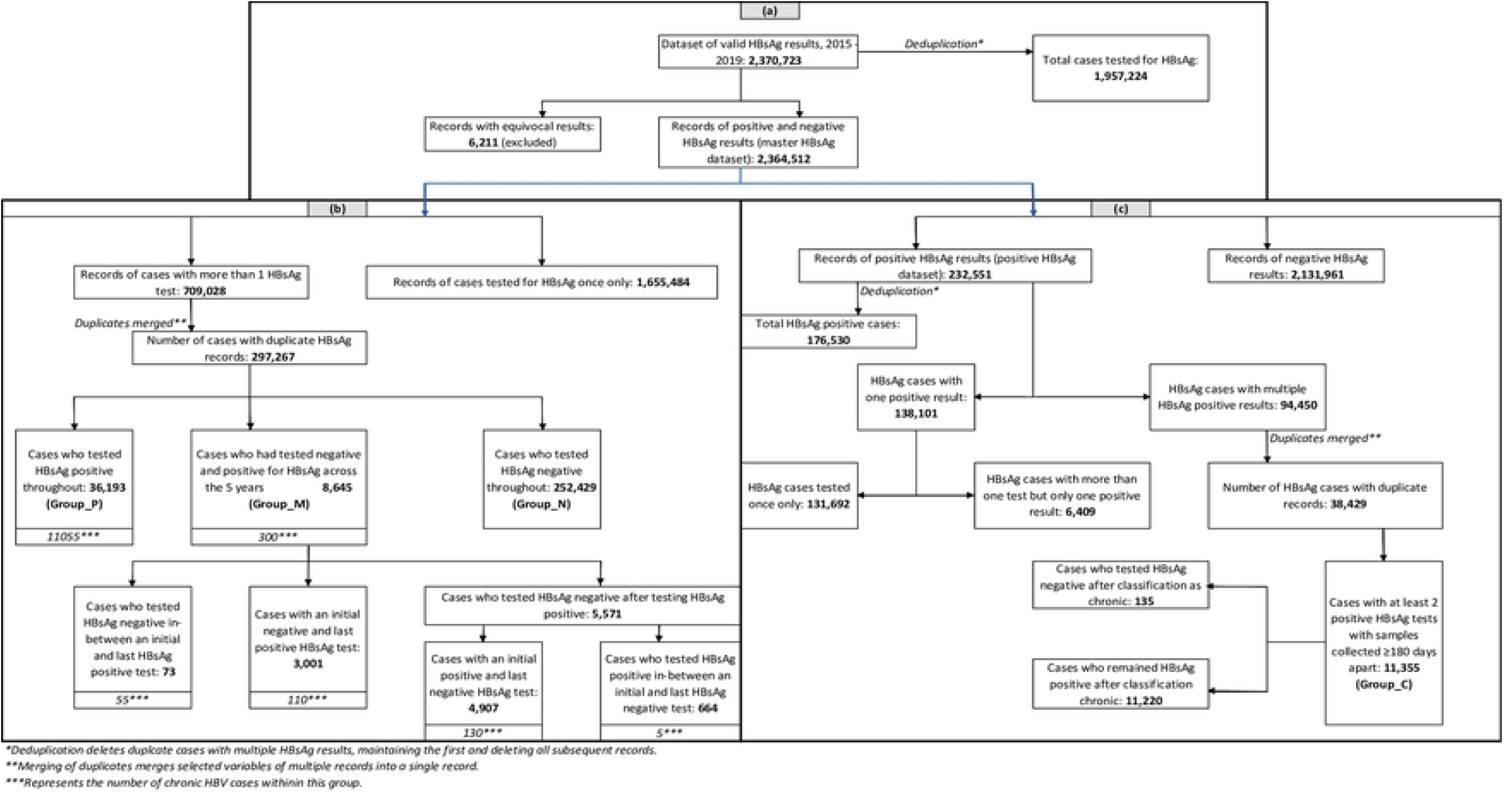
HBsAg sample number breakdown from the appended dataset of records from 2015 to 2019; from the dataset of valid HBsAg results to the master HBsAg dataset (a), from the master HBsAg dataset to a breakdown of Group_M (b), and from the master HBsAg dataset to a breakdown of Group_C (c)

### 3.1 Registry of cases who tested positive for HBsAg, 2015 to 2019

From the master HBsAg dataset of 2,364,512 records of positive and negative HBsAg tests, we identified and extracted 232,551 records of positive HBsAg results (positive HBsAg dataset) (Fig 1c). Deduplication of cases resulted in a line list (registry) of 176,530 cases who had tested positive for HBsAg at least once during the five-year period, with a test positivity rate of 9.0% (176,530 / 1,957,224) (Fig 1c).

#### 3.1.1 Analyses of the 2015 to 2019 HBsAg registry

Of the 176,530 HBsAg positive cases, significantly more males (94,698, 53.6%) tested positive than females (78,934, 44.7%, p < 0.0001) (Fig 2a and S2 Table). Individuals aged 25 to 44 years old had the highest proportion of HBsAg positive tests (115,906, 65.7%) (Fig 2b and S2 Table). Individuals 45 years and older accounted for 21.1% of the total HBsAg positive cases (37,200/176,530). The number of HBsAg positive cases 0 to 19 years old (4,980) was significantly lower than the number of HBsAg positive cases 20 years and older (166,259, p < 0.0001) (S2 Table). Amongst the 4,980 HBsAg positive cases 0 to 19 years old, 1,131 (22.7%) were under 5 years old (S2 Table). In the under 5-year age group, the HBsAg test positivity rate was 4.8%, and the HBsAg population positivity rate was 0.02% (Table 2).

**Figure 2:**
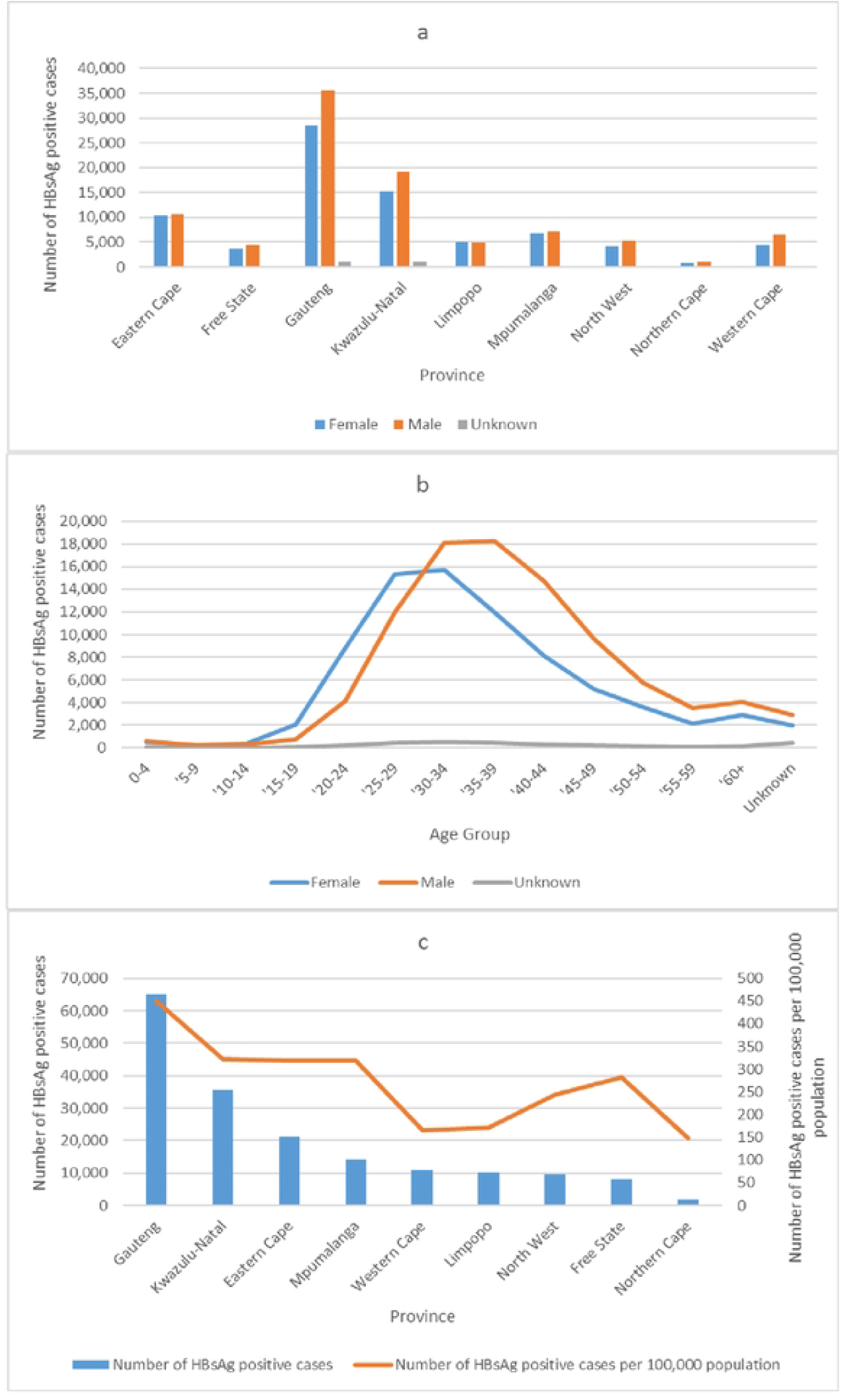
National distribution of HBsAg positive cases 2015 to 2019 (n = 176,530); province by gender (a) age group by gender (b), and province distribution versus 100,000 province population (c)

**Table 2:**
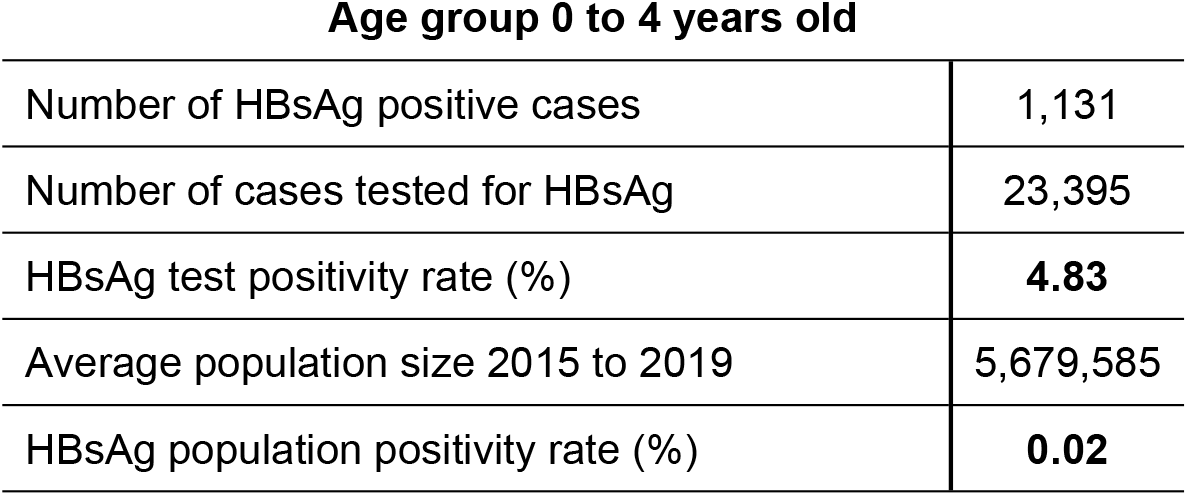
HBsAg test positivity and population positivity rate from 2015 to 2019.

| Age group 0 to 4 years old |  |
| --- | --- |
| Number of HBsAg positive cases | 1,131 |
| Number of cases tested for HBsAg | 23,395 |
| HBsAg test positivity rate (%) | **4.83** |
| Average population size 2015 to 2019 | 5,679,585 |
| HBsAg population positivity rate (%) | **0.02** |

At a provincial level, of the 176,530 HBsAg positive cases, Gauteng had the highest proportion of cases (65,085/176,530, 36.9%), followed by Kwazulu-Natal (35,532, 20.1%) and Eastern Cape (21,280, 12.1%), with the lowest proportion in the Northern Cape (1,827, 1.0%) (Fig 2c and S2 Table). Stratified by gender, the number of HBsAg positive males were significantly higher than females in eight of the nine provinces (p ≤ 0.0306), except in Limpopo with a significantly higher proportion in females (p = 0.0103) (S2 Table). The number of HBsAg positive cases per 100,000 population was highest in Gauteng (450/100,000), comparable between Kwazulu-Natal (322/100,000), Eastern Cape (318/100,000) and Mpumalanga (319/100,000), and least in Northern Cape (148/100,000) (Fig 2c).

### 3.2 Chronic HBV cases, 2015 to 2019

Of the positive HBsAg dataset of 232,551 records, we identified 138,101 cases who had only a single positive test over the 5-year period and 94,450 records of cases who had at least two positive HBsAg tests (Fig 1c). Of the 138,101 cases who had only a single positive test, 131,692 cases were tested once only over the five years and 6,409 cases were tested more than once but with only one positive test. On deleting the 138,101 cases and merging the duplicates of the 94,450 records, we remained with 38,429 cases with at least two positive HBsAg tests. Of the 38,429 cases, we identified 11,355 cases who had two positive tests with sample collection dates 180 days or more apart and classified them as HBV chronic cases, assigning them to Group_C (Fig 1c). Of the HBV chronic cases, 11,055 cases were from Group_P and 300 cases from Group_M (Fig 1b). Of the 300 cases in Group_M, 130 cases had an initial positive and last negative HBsAg test, 110 cases had an initial negative and last positive HBsAg test, 55 cases tested HBsAg negative in-between an initial and last positive HBsAg test, and 5 cases tested HBsAg positive in-between an initial and last negative HBsAg test (Fig 1b). One hundred and thirty five (135) cases tested HBsAg negative after being classified as chronic, whilst the majority (11,220) remained HBsAg positive (Fig 1c).

#### 3.2.1 Analyses of the chronic HBV cases, 2015 to 2019

Amongst the total HBsAg positive cases identified during the five-year period, 6.4% (11,355/176,530) were chronically infected. Analysis of the 11,355 HBV chronic cases showed a significantly higher proportion amongst males (6,211, 54.7%) than females (5,027, 44.3%, p < 0.0001), although at ages 15 to 29 years, significantly more females (1588, 62.8%) were identified as chronic carriers than males (941, 37%, p value <0.0001) (Fig 3a and S3 Table). The majority of the cases were amongst the ages of 25 to 49 years (9,230, 81.3%) (Fig 3b and S3 Table).

**Figure 3:**
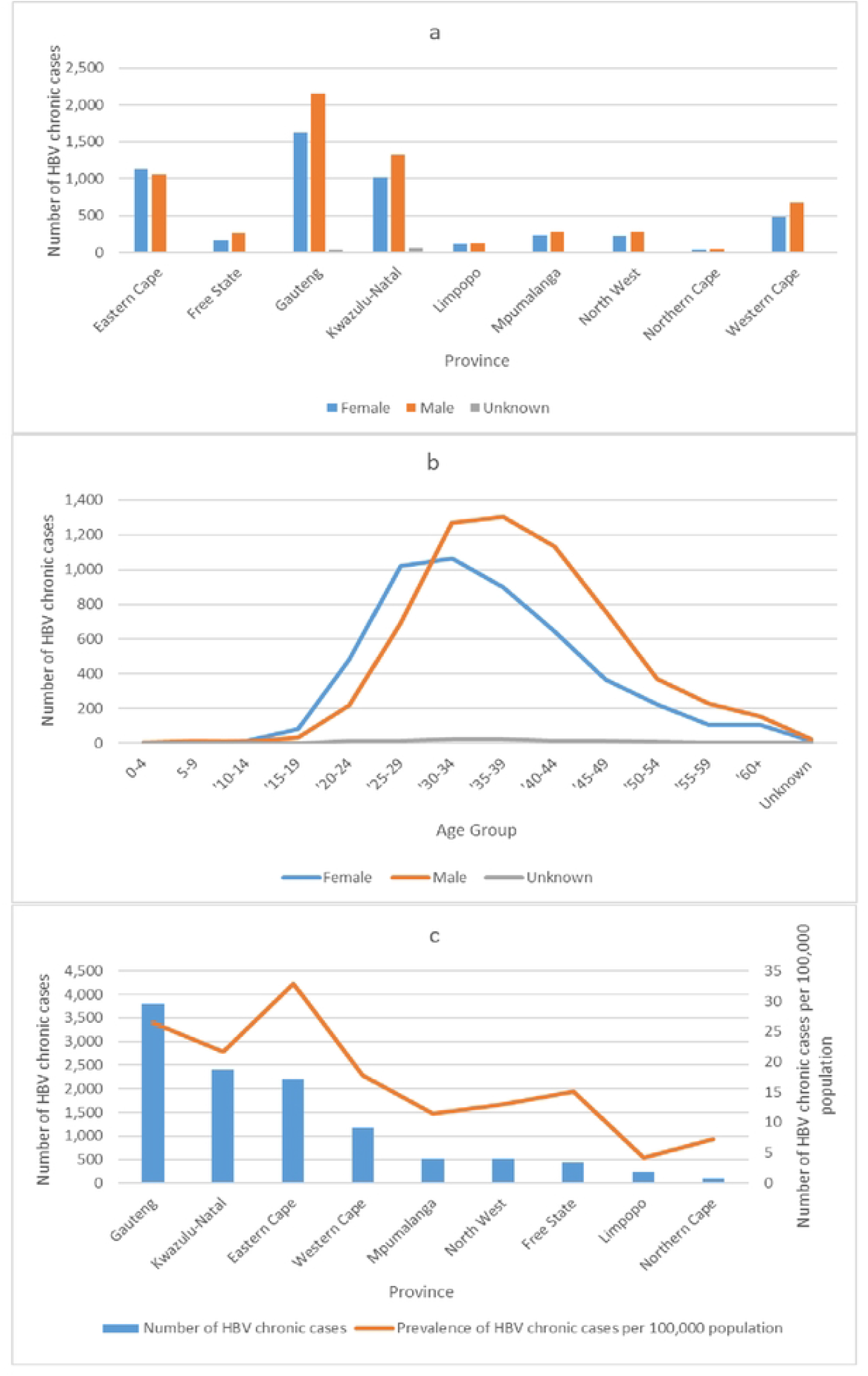
National distribution of HBV chronic cases 2015 to 2019 (n = 11,355); province by gender (a), age group by gender (b), and province distribution versus 100,000 province population (c)

Provincially, the majority of the chronic cases were from Gauteng (3,811, 33.6%), followed by Kwazulu-Natal (2,398, 21.1%) and Eastern Cape (2,198, 19.4%), with the least number of cases in Northern Cape (89, 0.8%) (Fig 3c and S3 Table). HBV chronic cases per 100,000 province population was highest in Eastern Cape (33/100,000), followed by Gauteng (26/100,000) and Kwazulu-Natal (22/100,000), and least in Limpopo (4/100,000) (Fig 3c). Stratified by gender, the number of HBV chronic males were higher than females in all provinces except Eastern cape, significantly higher in Free State, Gauteng, Kwazulu-Natal, Mpumalanga, North West and Western Cape (p ≤ 0.0254) (Fig 3c and S3 Table). In Eastern Cape, although more females were chronically infected than males, the difference was not significant (p = 0.1202) (Fig 3a and S3 Table).

## 4. Summary and Discussion

We analysed countrywide HBsAg laboratory data from 2015 to 2019 to determine the number of cases who tested positive for HBsAg and the number of cases with chronic HBV infection. Over the five years, the HBsAg test positivity rate was 9.0%, of which 6.4% were identified as chronic carriers. We noted that the majority of the HBsAg positive cases and HBV chronic cases were amongst cases who were born pre 1995, outside the vaccine-eligible era and therefore foreseeable. Numbers of HBsAg positive cases were much lower amongst individuals who were vaccine-eligible as infants (0 to 19 years), demonstrating effective vaccination in these groups. As vaccination is key to prevention of HBV infection, HBsAg positivity amongst vaccine-eligible individuals is likely due to the suboptimal vaccine coverage rates reported for South Africa from 2000 to 2019 (25-29). Suboptimal vaccine coverage rates may have largely been due to the challenges that faced EPI activities in South Africa, highlighted by EPI managers as being insufficient knowledge of vaccines and EPI practices among staff, financial constraints, staff shortages and high staff turn-over (43).

More males were HBsAg positive and chronically infected overall, although more females were HBsAg positive at ages 10 to 29 years and chronically infected at ages 15 to 29 years. The higher rate amongst males overall is compounded by the fact that the average population size over the five years was higher in females. The higher rates observed in females 10 to 29 years old are likely due to their increased risk of symptomatic infection following viral infections, coupled with higher health seeking tendency including antenatal visits at these ages (44). The higher rates in males overall and at later ages are likely due to the increased risk of persistent HBV infection in males resulting in their increased risk of HBV chronicity (45, 46). This is in keeping with studies conducted in Taiwan, Greece and New Zealand, where it was reported that females were less likely to develop chronic HBV infection than males (46-48). In a study on sex differences in response to HBV infection, Blumberg reports that the prevalence of chronic carriers amongst males is higher than females in most human populations (49). In children 0 to 4 years old, we noted a sizeable number of HBsAg positive cases from 2015 to 2019 (1,131). With no birth dose of the HBV vaccine administered to these children and no routine screening for HBsAg at antenatal care, we suspect a considerable proportion were infected from their mothers given the increased risk with no preventative interventions (6, 10, 23, 24). Although horizontal transmission has been reported as a major transmission route between children in South Africa, justifiable reasons are lacking (20, 50). Considering the WHO impact target of ≤0.1% HBsAg prevalence for measuring elimination in children under 5 years old and the South African under 5 year HBsAg population positivity rate (population prevalence), South Africa is well below the target over 2015 to 2019 at 0.02% (14). However, the data presented are only from children tested in public healthcare facilities and it is likely that a substantial proportion of children under 5 years old were not HBsAg diagnosed given the low risk of symptomatic HBV infection in children (1). Substituting the denominator with the total number of cases under 5 years old who were tested for HBsAg over the five years, the HBsAg test positivity rate (test prevalence) was 4.83%, well above the 0.1% target.

Provincially, we noted that the number of HBsAg positive cases and the number of HBV chronic cases were highest in Gauteng, followed by Kwazulu-Natal and Eastern Cape and lowest in the Northern Cape, not surprising given that the average population size of these four provinces from 2015 to 2019 follows the same pattern. In addition, the findings in Gauteng and Northern Cape may also be related to the land size of these provinces versus their population sizes (Table 1) (51). Furthermore, Northern Cape boasted the highest average vaccine coverage rate from 2000 to 2019 (85.57%) (S1 Table). Taking into consideration population size per province, Eastern Cape was seen to have the highest number of chronic HBV cases in the country (33/100,000), in keeping with one of the poorest vaccine coverage rates averaging 69,54% from 2000 to 2019. Surprisingly, however, there were more chronic infections in females in Eastern Cape, despite significantly higher HBsAg positive tests in males. Regarding gender distribution, we saw that significantly more females tested positive for HBsAg in Limpopo province, yet more males were chronically infected. We link this finding in Limpopo to the increased risk of symptomatic infection following viral infections in females together with the increased risk of persistent HBV infection in males (44-46). Although reasons are unclear, we are cognisant of provincial public health disparities within South Africa and suboptimal vaccine coverage rates (S1 Table) (52). Therefore, provincial investigations, including community awareness, HBV testing and follow up practises, social practises, and vaccination policies would provide insight into provincial HBV burden.

From analyses of cases who tested negative and positive for HBsAg over the five years (Group_M), we saw that 64.4% (5,571/8,645) of cases tested negative after a positive test, suggesting HBsAg clearance and resolution of HBV infection. Analyses of testing patterns showed that amongst the cases who tested negative after a positive test, 11.9% (664/5,571) of cases tested negative before testing positive, indicating that in these individuals HBV infection was likely acquired during 2015 to 2019 and likely amongst those individuals who did receive or complete their primary course of vaccinations. We also observed a small proportion (73/8,645, 0.8%) of cases from Group_M who tested HBsAg negative between two positive tests, suggesting HBsAg clearance after acquiring HBV infection followed by reactivation of HBV disease or HBV reinfection, a phenomenon seen amongst HIV-1 infected individuals (8). Amongst the 11,355 chronic HBV cases, we report a last negative HBsAg test over the five years in 1.2% of cases following classification as chronic, in line with reported statistics of HBsAg clearance observed in 1-2% of chronic infections following the natural course of disease or treatment interventions (18, 19).

A limitation of this study is that we report findings restricted to passive surveillance over a fixed period of time. Population prevalence rates include South Africa’s total population as the denominator and our findings are from individuals attending only public healthcare facilities. Our numbers, therefore, represent minimum estimates. Our findings of provincial differences may represent provincial disparities rather than disease burden. In addition, the province is allocated based on the location of the testing facility and patients may have travelled to facilities in provinces outside their place of residence. HBsAg positivity amongst children 0 to 4 years old may be associated with transient positivity soon after vaccination, although reasons for testing in this age group are unclear. Our data were not linked to HIV status and we therefore cannot report on HBV and HIV co-infection or relate our findings to other studies reporting HBV prevalence and chronicity in HIV-infected individuals. A strength of our study, however, is the interrogation of a robust nationally representative public health dataset comprising over 2,3 million records.

## 5. Conclusion

Prevention of HBV infection through infant vaccination, inclusive of a birth dose, is key to eliminating HBV infection. Given that the HBV vaccine is administered as a hexavalent vaccine in South Africa, optimal (3-dose) HBV vaccine coverage would not only reduce HBV burden, but would provide protection from other debilitating vaccine-preventable diseases. Early identification of HBV chronicity is fundamental in reducing the risk of liver cirrhosis and hepatocellular carcinoma through appropriate treatment initiation, and can be achieved through real-time data analysis and dissemination of the relevant information for public health interventions.

## Data Availability

Data used for this study has been uploaded as three supporting information zipped files.

## Acknowledgements

We acknowledge the NHLS CDW department for providing the 2015 to 2019 hepatitis B data.

## Supporting information

**S1 Table: Vaccination coverage amongst all children under one year who have completed their primary course of immunisation in South Africa**

**S2 Table: Distribution of HBsAg positive cases by province and age group stratified by gender**

**S3 Table: Distribution of HBV chronic cases by province and age group stratified by gender**

